# Sensitivity of temperature-based time since death estimation on measurement location

**DOI:** 10.1101/2023.04.18.23288727

**Authors:** J. Shanmugam Subramaniam, M. Hubig, H. Muggenthaler, S. Schenkl, J. Ullrich, G. Pourtier, M. Weiser, G. Mall

## Abstract

Rectal temperature measurement (RTM) from crime scenes is an important parameter for temperature-based time of death estimation (TDE). Various influential variables exist in TDE methods like the uncertainty in thermal and environmental parameters. Although RTM depends in particular on the location of measurement position, this relationship has never been investigated separately. The presented study fills this gap using Finite Element (FE) simulations of body cooling. A manually meshed coarse human FE model and an FE geometry model developed from the CT scan of a male corpse are used for TDE sensitivity analysis. The coarse model is considered with and without a support structure of moist soil. As there is no clear definition of ideal rectal temperature measurement location for TDE, possible variations in RTM location (RTML) are considered based on anatomy and forensic practice. The maximum variation of TDE caused by RTML changes is investigated via FE simulation. Moreover, the influence of ambient temperature, of FE model change and of the models positioning on a wet soil underground are also discussed. As a general outcome, we notice that maximum TDE deviations of up to ca. 2-3 h due to RTML deviations have to be expected. The direction of maximum influence of RTML change on TDE generally was on the line caudal to cranial.

## Introduction

Temperature based death time estimation (TDE) is crucial in homicide investigations. TDE is estimated from core temperature measurement data using the model curve T(t) comprising a priori knowledge of the postmortem rectal temperature decline. The phenomenological approach and the physics-based approach are two different techniques used to generate the model curve T(t). The method of Marshall and Hoare with the Henßge parameters from the Nomogram method (MHH) is a prominent phenomenological approach using a double exponential model with fitted parameters for modeling rectal cooling [1]. Physics-based approaches use the heat transfer equation considering heat exchange mechanisms, cooling conditions and thermal material properties. Finite element (FE) based TDE method (FEM) is a physics-based approach with reasonable computational effort (see e.g. [2, 3, 4]). For an extensive overview over temperature based TDE see e.g. [5].

In any TDE method, error quantification is a desideratum as TDE results can lead to acquittal or conviction of suspects. Three different errors exist in FEM such as errors due to space and time discretization, input data errors, and model errors. They lead to the deviation of the TDE value from the actual value. Though several approaches for error estimation exist for TDE e.g. for MHH [6] and for FEM [7] none of them takes into account the uncertainty introduced in TDE due to input data errors caused by variations in the rectal temperature measurement location. The present article tries to close this gap using an FE approach. This does not, however, limit the results to FE TDE methods.

Rectal temperature measurement (RTM) can be performed using approved devices and without the necessity to injure the body. However, there is a certain amount of diversity in the characterization of the measurement locus in forensic literature. In early well-known TDE studies [8], [9], it is reported a 3 – 4 inches [7 – 10 cm] insertion depth for RTM. Marshall and Hoare [10] mention the relationship between the liver and rectum temperature. Henßge [11, 12] states that the insertion depth for RTM should be at least 8 cm from the musculus sphincter ani. In [11] Henßge estimates the precision of rectal temperature measurement to be about “+-2 °C”. Further, Henßge [13] emphasized the usage of linear, rigid and not very flexible thermometers to achieve an insertion depth without applying force. He also recommends a temperature measurement at the mesenteric root in the lower left abdomen additional to the temperature measurement in the deep abdominal space [14]. Moreover, Henßge advises the thermometer to be inserted in the rectum as deep as possible without applying any force [15]. Indeed, a lot of discrepancy in rectal temperature measurement location exists in literature, so it is necessary to study its effects on TDE. Hence, this motivates a sensitivity analysis of TDE with respect to measurement locus.

The study was executed with participation of the Institute of Legal Medicine (IRM) at the University Hospital Jena of the Friedrich-Schiller-University Jena and of the Zuse Institute Berlin (ZIB), an interdisciplinary research institute for applied mathematics and data-intensive high-performance computing.

Additional computation results are presented in the Supplementary Information (SI) associated with the electronic edition of the article available via the website of the journal.

## Method

### Finite Element model

Background information on numerically solving partial differential equations and on FE simulation can be found e.g. in [16].

For one exemplary case of human corpse with a given CT-scan, two different methods were used to generate FE models. In one of the methods, the FE Model (CTM) was developed from the segmentation of a CT scanned human body and in the other, the human body was manually approximated by hexahedral elements (see e.g. [2], [17]). Two variants of this manually generated model were constructed: One model (CM) is floating freely in air and a copy (CMS) of CM is laying firmly on a wet soil substrate. Detailed simulations of physical heat transfer processes generated the rectal temperature curves T(t). The corpse cooling was computed in all of the FE models based on the well known heat transfer equation (1), a partial differential equation where c is the specific heat capacity, ρ the mass density, and κ the heat conductivity of the tissue:

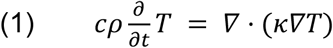

Heat transfer from the body to the environment across the skin due to convection and surface-to-ambient radiation was captured by a Robin boundary condition with effective heat transfer coefficient γ as given in equations (2a), (2b) (see e.g. [7]):

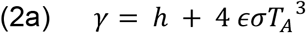

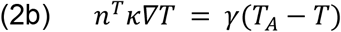

Here, T_A_ is the ambient temperature, h is the heat transfer coefficient, ε is the emissivity and σ is the Stefan-Boltzmann constant. In the CTM developed from the CT scan of a human body, the initial temperature field T_0_ defined at time t = 0 satisfies Pennes’ Bio-Heat-Transfer-Equation (BHTE) [18]

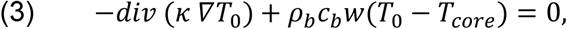

where ρ_b_c_b_ is the heat capacity of blood, w is tissue perfusion, and T_core_ is the body core temperature.

In the manually developed FE model CM, the initial temperature field T_0_ is defined with a gradient between core and outside elements as in [2] referring to physiology literature.

For reliable comparison between the different models, the equivalent effective heat transfer coefficient was applied to the CTM corresponding to the convection and radiation terms applied on CMS and CM. The model curves T(t) were sampled at the designated nodes C and SP_R_k (k = 1,…6) corresponding to the anatomical RTM locations (RTML). The point C was placed at the intended measurement location in the respective FE model, whereas the six points SP_R_i (i=1,…,6) lay on the vertices of an octahedron with radius R. The SP_R_i were defined to represent possible locations of a misplaced temperature sensor. If no confusion can arise about the value of R the SP_R_i are abbreviated SPi.

Our results consist of maximum deviations D_MAX_^M^ comparing two cooling curves computed at two locations of RTM’s in a specified FE model M = CTM, CM, CMS. The distance D ^M^ is defined between the cooling curves T_C_(t) and T_SPi_(t) at the center point C of a measurement point octahedron (see Fig. 4) of radius R and at its vertices SP_R_k (k = 1, …, 6). We also computed differences D_MAX_^M1,M2^ := D_MAX_^M1^ – D_MAX_^M2^ of those maximum deviations for two pairs (CM-CMS and CM-CTM) of different FE models (M1, M2) for all C – SP_R_i pairs. All computations were performed for different constant ambient temperatures T_A_ and for different octahedra radii R. We also performed all of the computations for each of four Q-ranges, where the first three are Henßges Q-ranges (see e.g. [15]).

### CT meshed FE model CTM

All of the modeling work done on the CT meshed FE model CTM was performed at the Zuse Institute Berlin (ZIB). The CT scan of a male corpse of length L = 1.74 m and weight M = 62 kg was segmented in the software AMIRA and the segmented data was converted into an FE model, comprising of 961234 tetrahedral elements, via the FE-program Kaskade (see e.g. [19]), an in-house code of ZIB. As depicted in Fig. 1, the feet of the corpse were not included in the model and the weight, size and volume of the whole model were scaled according to the scanned corpse dimensions. The segmentation of different body parts was carried out using differences in density and location of different tissue types like bone, fat, bladder, kidney, abdomen, liver, heart, lungs and muscle as seen in Tab. 1 in the Appendix.. In CTM, skin tissue was not segmented separately from the subcutaneous fat tissue since skin tissue was assumed to have a negligible effect on the TDE. Hence, the skin material properties were not considered in CTM.

**Fig. 1.**
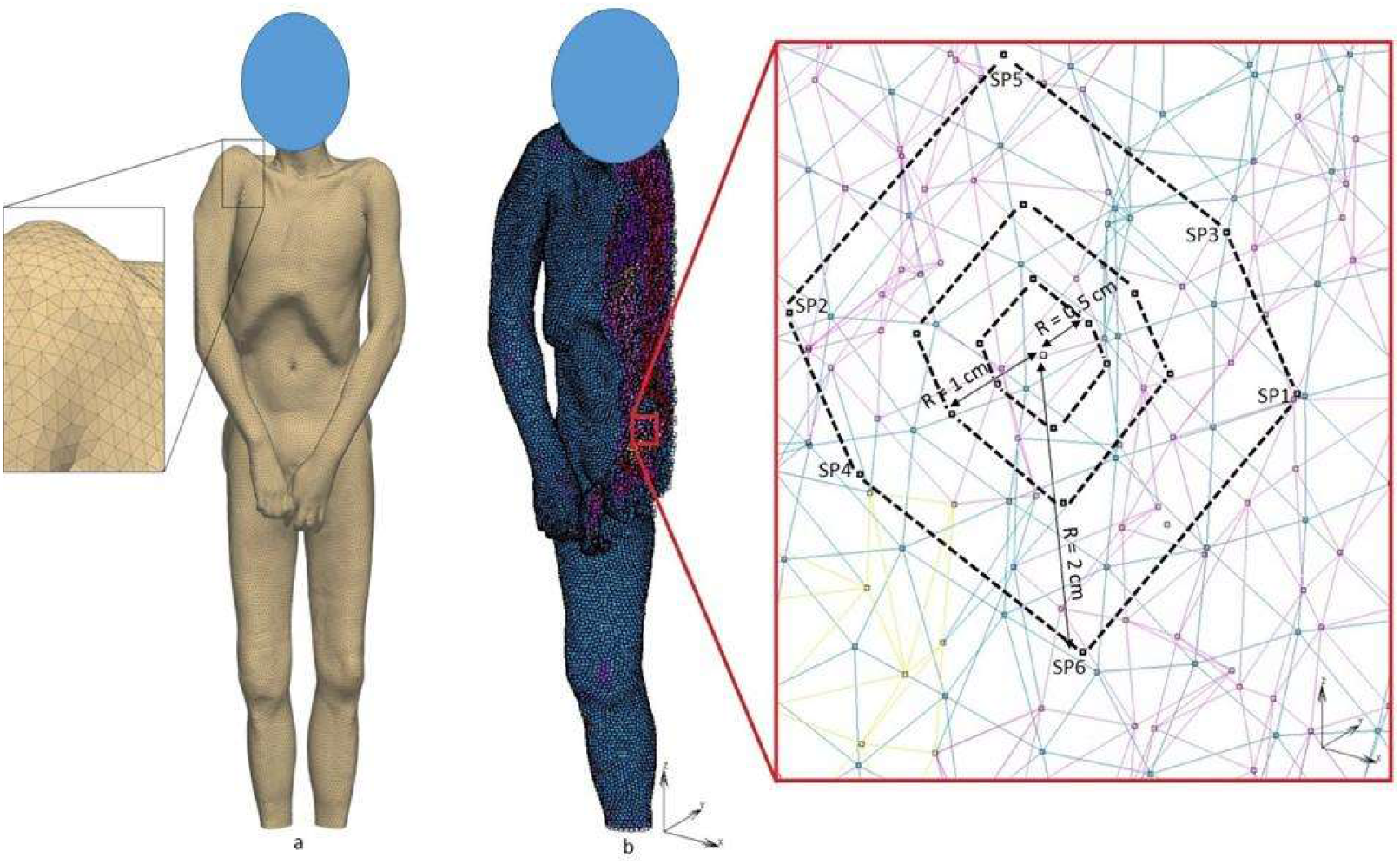
**a:** CT Meshed male corpse model with magnified mesh detail. **Fig. 1 b:** Right half view of CTM model with magnified measurement location showing measurement points C, SPi (i = 1,…,6) in three different octahedra of radii R = 0.5 cm, 1 cm, 2 cm. For the octahedra see Fig. 4. Only the contour lines of the octahedra are shown for clarity.

### Coarse meshed FE model CM

Our coarse meshed FE model CM was already described in several publications [2, 3, 20, 21]. Its mesh was manually constructed using the FE preprocessor Mentat. It consists of approx. 12000 nodes in approx. 8300 hexahedral elements with trilinear shape functions. The model contains numerous compartments (see Fig. 2) standing for distinct anatomical structures of the body.

**Fig. 2.**
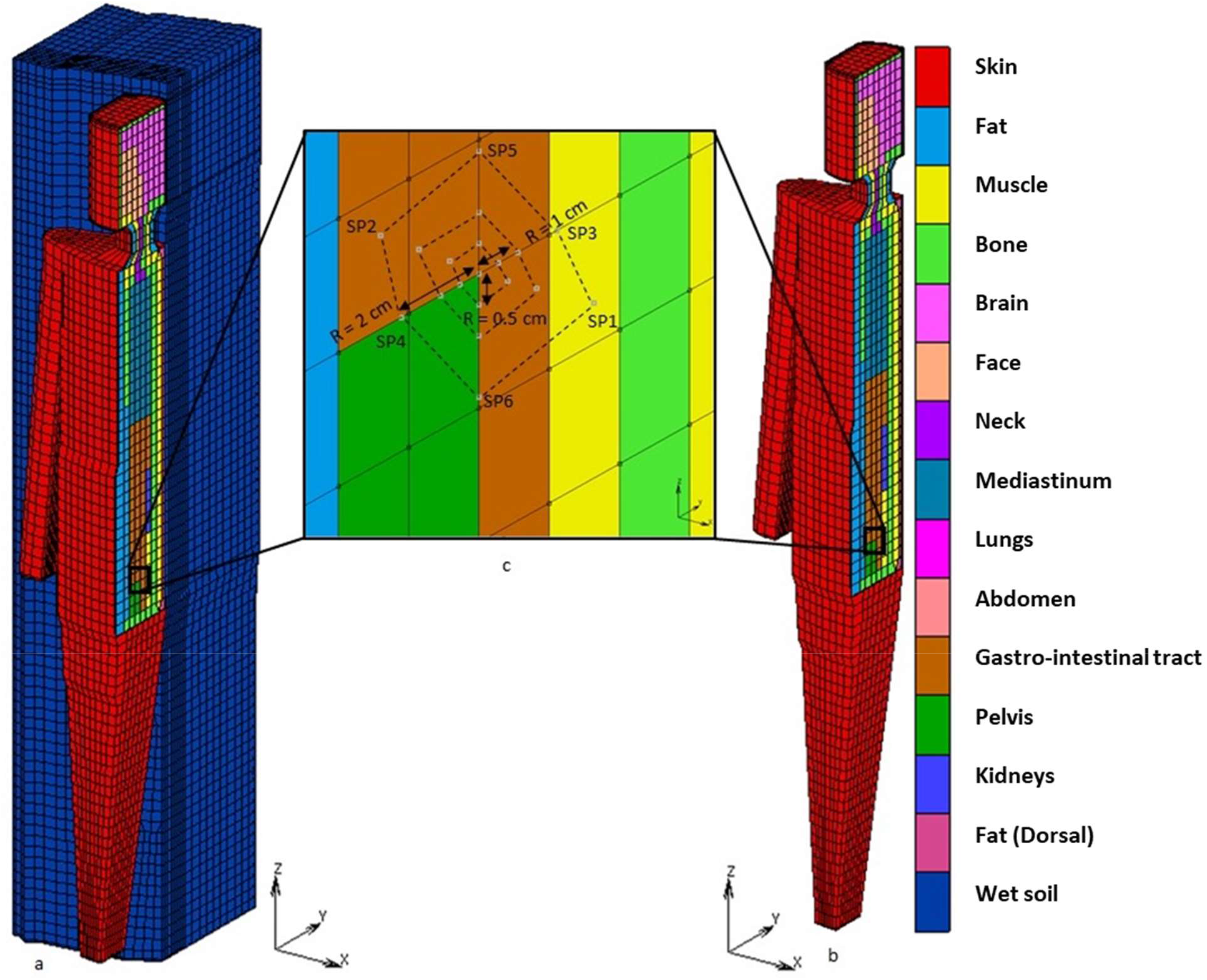
**a:** Coarse model lying on a support structure (CMS), **Fig. 2 b:** Coarse model without support structure (CM), **Fig. 2 c:** Detailed view of measurement points C, SP_1_,….,SP_6_ along different radii R = 0.5 cm, 1 cm, 2 cm. For the octahedra see Fig.4. Only the contour lines of the octahedra are shown for clarity.

**Fig. 3:**
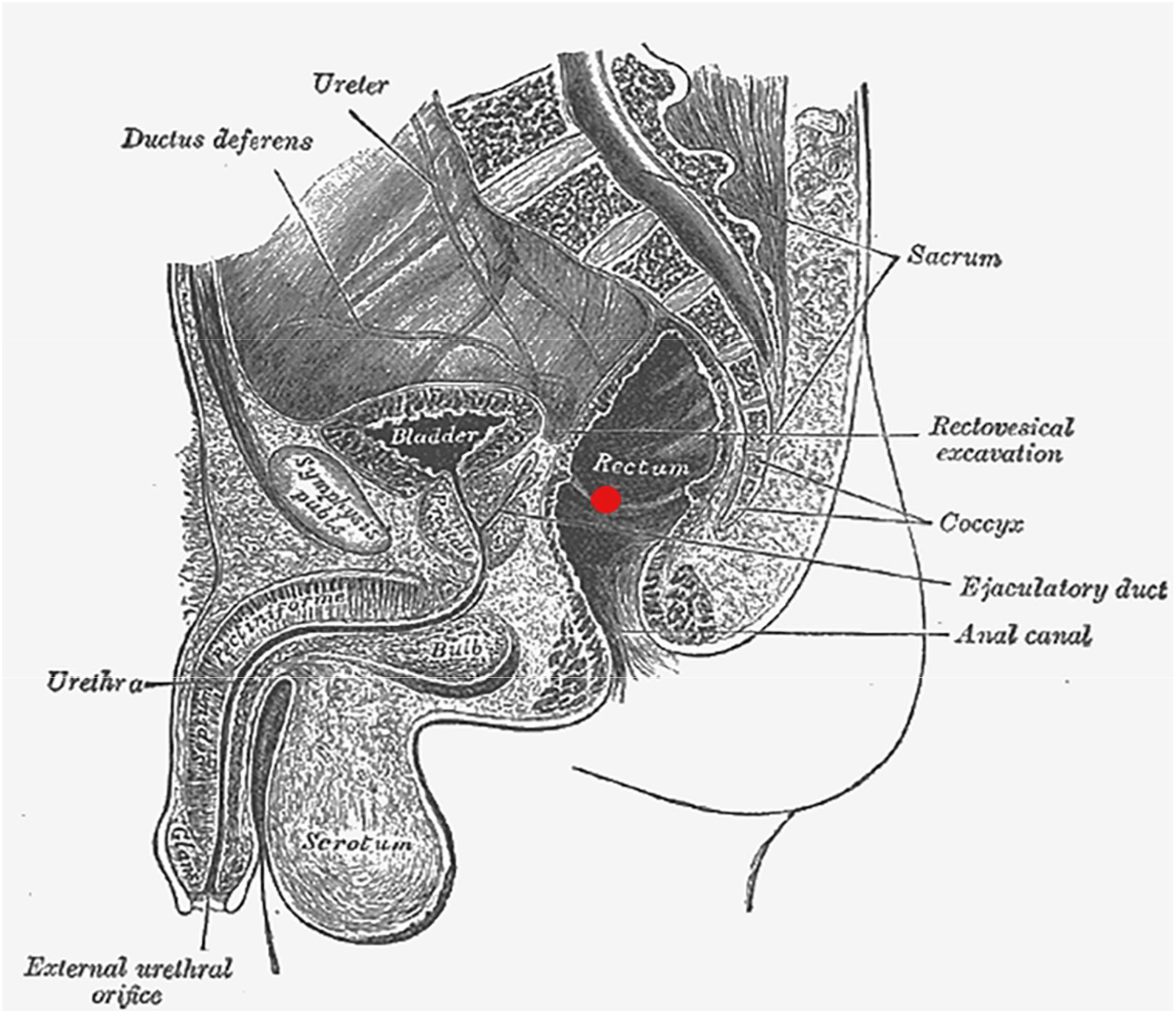
Anatomical sketch depicting ideal measurement location [23]

**Fig. 4:**
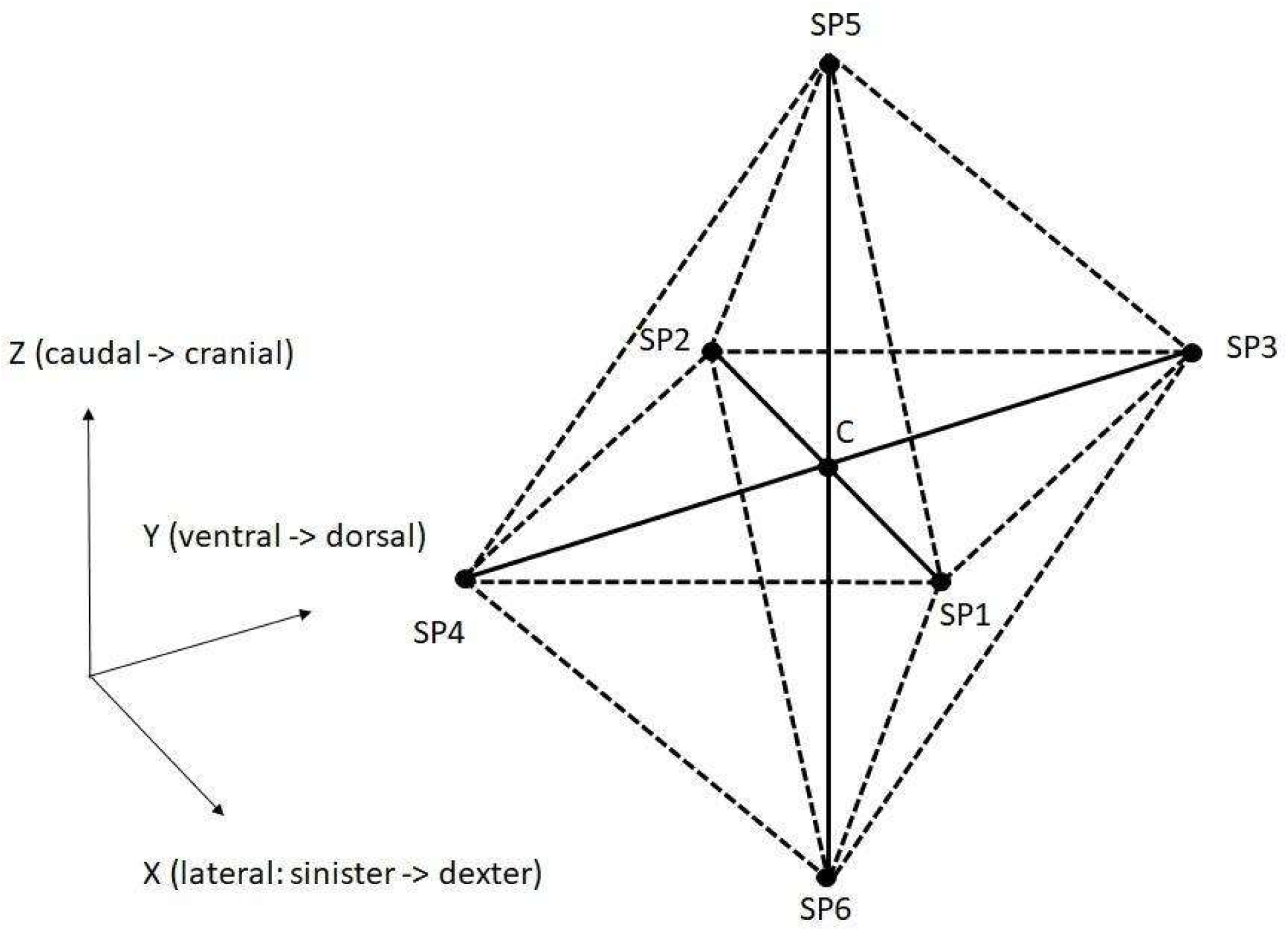
Central position C and six additional measurement positions (SP_R_k =) SPk (k = 1,…,6) in X, Y and Z direction in CTM, CMS, CM model at different radii R = 0.5 cm, 1 cm, 2 cm from center position C.

Our standard FE model of length L = 1.64 m and mass M = 64 kg was scaled geometrically to a length of L’ = 1.74 m and weight of M’ = 62 kg, corresponding to the male reference body of our study. The scaling factors are defined as in the equations (4) and (5) where k_1_ is linear 1-dim scaling along the body length axis and k_2_ is linear 2-dim dilation in the transverse plane [2].

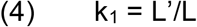

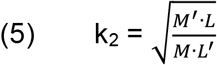

Generation and scaling of FE mesh and post-processing were performed in MSC’s pre- and post-processing tool Mentat.

Further, to investigate the TDE sensitivity on more complex boundary conditions favoring thermal energy transfer from the body core in dorsal direction, a supporting structure or floor is modeled with an hexagonal FE mesh as shown in Fig. 2 a. The FE model lies on its back on the support structure defined, presumably causing large differences between C-temperature and temperature at SP4 or SP3, i.e. along y direction. The support structure is defined with the thermal properties of wet soil such as thermal conductivity c = 2 W/m^2^K, specific heat κ = 2200 J/kgK, density ρ = 1900 kg/m^3^ [22] and emissivity ε = 0.95. The coarse model laying on its back on the support structure is abbreviated CMS, whereas the coarse model without support structure is named CM (see Fig. 2 c). CMS and CM were scaled to the same height and weight as CTM. This will eliminate the influence of weight on the body cooling.

### Definition of measurement positions C and SP_**R**_**k**

TDE estimations are usually based on a single RTM. Hence, the measurement position is subject to variations due to different anatomies, thermometer angles and insertion depth. An approximate ideal measurement position can be specified from anatomy including forensic knowledge on measurement locations used in practice. The central position C for our measurements was determined in the pars ampullaris of the rectum near the incisura transversalis, which is the passage from pars ampullaris to the pars sacralis of the rectum.

Additional measurement positions SP_R_k (k = 1, …, 6) for our sensitivity studies were chosen in spatial x, y and z direction at distances (octahedral radii) R = 0.5 cm, 1 cm, 2 cm from the central measurement position C. For each fixed distance R six additional measurement locations SP_R_k were established. The six additional measurement positions SP_R_k form an octahedron with the central point C in its middle. The schematic sketch representing the additional positions in the model was illustrated in Fig. 4.

The anatomical position of additional measurement points in CTM are described as follows: SP1 lies in the center of abdomen surrounded by abdomen tissues, SP3 lies nearby bone, SP4 lies in the center of abdomen, SP6 lies very near to abdomen. Fig. 1 b shows the anatomical positions of measurement points in CTM. Similarly, the anatomical position of additional measurement points in CMS and CM are depicted in Fig. 2 c, which helps to identify the position of additional measurement points. SP1 lies lateral from C in positive X direction. Due to CM’s left-right symmetry the temperature value at SP2 is equal to the temperature at SP1. The points SP2, SP3, SP5 lie in the gastrointestinal tract. SP6 and SP4 lie on the border between gastrointestinal tract and pelvis.

### Simulation

The CT-generated model CTM was imported from the FE research code Kaskade 7 [19] into the commercial MSC-Marc Mentat FE system and the material parameters are defined as given in Table 1 of [2] (see Tab. 1 in the Appendix). The same material properties were used also for CMS and CM models. The initial temperature field T_0_(r) at the time of death on the body was calculated via the Pennes’ Bio-Heat-Transfer-Equation BHTE on the original CTM model [18]. The field T_0_(r) was approximated by 10 discrete node sets corresponding to 10 discrete temperatures in the converted version of CTM. In the CMS and CM models, the initial temperature was defined according to [2] where it was taken from physiological textbooks. The simulation is carried out for three different constant ambient temperatures T_A_ = 5° C, 15° C, 25° C. The convection and radiation parameters in CTM were defined by the effective heat transfer coefficient γ = 4·ε·σ T_A_^3^ [7] and the calculated values in increasing order of T_A_ = 5° C, 15° C, 25° C are γ = 7.93 W/m^2^K, 8.45 W/m^2^K, 9 W/m^2^K. In CMS and CM, a convection coefficient of h = 3.3 W/m^2^ K is applied [2]. Radiation view factors in the CMS and CM models were computed by Monte-Carlo-simulation [2]. Although the method of defining boundary conditions differs between CTM and CM models, it was found that the linearization of the radiation term applied in CTM has negligible effect on the cooling curve [7]. A sparse direct algorithm in MSC-Marc 2020 and adaptive time stepping with maximum allowed temperature change of 1°K and 0.1s initial time step were used.

**Tab. 1:** Material properties used in CTM, CM and CMS model [Mall 2005A]

|  | CTM, CM and CMS model |  |  |
| --- | --- | --- | --- |
| Property | Conductivity | Capacity | Density |
| Unit | W/mK | J/kgK | Kg/m<sup>3</sup> |
| Skin | 0.47 | 3680 | 1085 |
| Fat | 0.21 | 2300 | 920 |
| Muscle | 0.51 | 3800 | 1085 |
| Bone | 0.75 | 1700 | 1357 |
| Brain | 0.49 | 3850 | 1080 |
| Face | 0.51 | 3245 | 1056 |
| Neck | 0.48 | 3363 | 1006 |
| Median intestinal Organs | 0.47 | 3375 | 1033 |
| Lungs | 0.28 | 3520 | 560 |
| Stomach | 0.48 | 3730 | 1080 |
| Abdomen | 0.46 | 3346 | 933 |
| Bladder | 0.49 | 3350 | 1008 |
| Kidney | 0.39 | 3158 | 1026 |
| Fat back | 0.21 | 2300 | 920 |
| Wet soil | 2 | 2200 | 1900 |

### Quantification of TDE deviations

TDE deviation from the central position C to any position SP_R_k is quantified by the maximum distance DMAX between the respective cooling curves, which is defined in this section. Let the time interval [a, b] be given during body cooling which starts at 0 h and let a = 1 h be the left point and b = 45 h be the right point of our interval. Let further T_1_(t) and T_2_(t) be two cooling curves defined on the interval [0, b]. Evaluation of the cooling curve distances started at a > 0 h for reasons of numerical stability. The ambient temperature T_A_ is assumed constant in space and time outside the body. The ***reference curve*** T_R_(t) for two cooling curves T_1_(t) and T_2_(t) is constructed as the pointwise mean of T_1_(t) and T_2_(t):

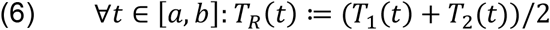

Let T_0_ := T_R_(0) be the initial temperature at the center point C. Now the reference curve T_R_(t) can be normalized to the function Q(t) taking values in the real number’s interval [0, 1]:

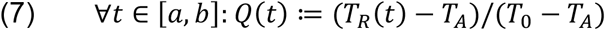

This makes it possible to define the Q-intervals Q_1_ to Q_4_ (where Q_1_, Q_2_, Q_3_ correspond to Henßge’s [15] normed temperature ranges for tolerance radii) and their left and right boundaries q_0_, …, q_4_ by:

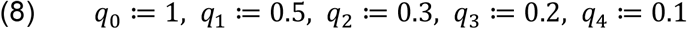

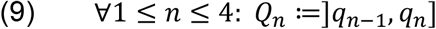

Assuming T_R_(t) to be strictly monotonically decreasing, we can uniquely map the Q-interval boundaries q_n_ to the respective temperature values T_R(4-n)_.

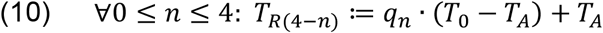

Since the two cooling curves T_1_ and T_2_ are monotonically decreasing it is possible to define their inverse curves t_1_, t_2_, called the **(absolute) time since death (TSD) estimation result curves** w.r.t. the cooling curves T_1_, T_2_ on the domain [T_MIN_,T_MAX_] which is the range of the reference curve T_R_ on the temperature axis by:

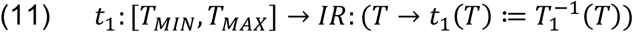

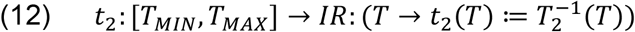

The time functions t_1_, t_2_ are frequently concatenated with the reference curve T_R_, thus giving t_1_(T_R_(t)), t_2_(T_R_(t)) in the following definitions. For short those concatenations will be abbreviated t_1_(t), t_2_(t). If the point t in time is clear, we will even write t_1_, t_2_. We will now quantify the distance between the cooling curves T_1_ and T_2_, which are actually constructed as distances between the inverse curves t_1_ and t_2_. The reason for this construction lies in our primal interest in time differences because our research’s number one target is *time since death*. All of the temperature curve distance measures are constructed using the (time oriented!) maximum distance D_MAX_(T_1_,T_2_) between two cooling curves T_1_ and T_2_, which is shown here for continuous functions first. The maximum domain of the reference curve T_R_ on the time axis is the interval [t_MIN_,t_MAX_] which is taken therefore as the domain for maximum-finding in our definition :

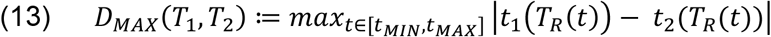

We will now consider the equivalent of the definition (13) in terms of real measured temperature curves T_i_(t). Each curve T_i_(t) is represented as a finite series of real measurement values (T_i_^1^, …, T_i_^N^) on a finite (regular) grid of time values (t^1^, …, t^N^) which is the same for both of the curves. The first point t^1^ of the time grid is identical to the starting time of cooling computation, while the last point t^N^ marks the end of the cooling computation interval. To provide a joint domain of definition for both our inverted temperature curves T_R_^-1^ we define:

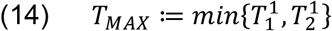

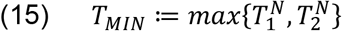

Now we have constructed the range of our reference curve T_R_(t). The limits of its domain of definition [t_MIN_,t_MAX_] can then be computed easily:

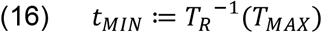

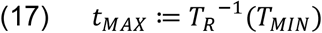

Let K be a natural number and let (t^1^, …, t^K^) be a regular time grid on the interval [t_MIN_,t_MAX_] with t^1^ := t_MIN_ and t^K^ := t_MAX_. This provides the equidistant sampling points for the final definition of the functions t_i_(T_R_(t)). Let further be (T^1^, …, T^K^) the corresponding temperature grid with T^1^≤ … ≤ T^K^ and T^k^ := T_R_(t^k^) for all k = 1, …, K and T^K^ := T_MAX_. The time grid’s width Δt is:

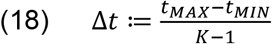

Let further be t_qn_ the inverse image of T_Rn_ under the reference function T_R_ for all n = 0, …, 4:

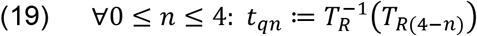

The five points t_qn_ on the time scale constitute the endpoints of four time intervals t_Q1_, …, t_Q4_:

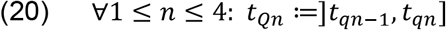

We will now redefine our measure (13) quantifying the distance between the cooling curves T_1_ and T_2_ in terms of the real samples (t^1^, …, t^K^) and (T_i_^1^, …, T_i_^K^). The distance measure is the distance D_MAX_ (T_1_, T_2_). It is defined in a global version on the whole time interval [t_MIN_, t_MAX_] and in four local ones residing on one of the intersections t_Qj_ ∩ [t_MIN_, t_MAX_] on the time axis each. For all 1 ≤ i ≤ 4 the number of t^k^ lying in t_Qj_ ∩ [t_MIN_, t_MAX_] is denoted by K_i_.

The ***(absolute global) maximum distance*** D_MAX_(T_1_, T_2_) is defined as:

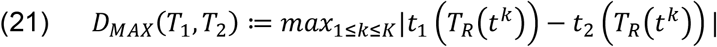

while the ***(absolute) Q***_***i***_***-local maximum distance*** D_MAX,Qi_(T_1_, T_2_) is defined for i = 1, …, 4:

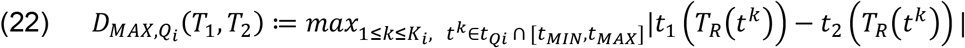

## Results

In a first step we evaluated the results for the direction in space (X,Y,Z as in Fig. 4), where the largest TDE-deviation value D_MAX_ caused by RTML -variation was seen. Interestingly, this *maximum direction depended neither on the ambient temperature T*_*A*_ *= 5 °C, 15 °C, 25 °C nor on the octahedron radius R = 0*.*5 cm, 1 cm, 2 cm*. The FE-model chosen makes the only difference. Therefore we get the following list of directions for maximum D_MAX_ caused by RTML-variation:

- CM: Z caudal - cranial
- CTM: Y ventral - dorsal
- CMS: Y ventral - dorsal

TDE deviations measured by D_MAX_^M^ on the FE model M = CTM, CMS, CM respectively depending on ambient temperature T_A_ and measurement radius R are represented in Fig. 5 – Fig. 7. The differences of TDE deviations D_MAX_^M1,M2^ on the model pairs (M1, M2) = (CM, CTM), (CM, CMS) as depicted in Fig. 8 – 9 were calculated as follows: D_MAX_^CM,CTM^ = D_MAX_^CM^ – D_MAX_^CTM^ and D_MAX_^CM,CMS^ = D_MAX_^CM^ - D_MAX_^CMS^.

**Fig. 5:**
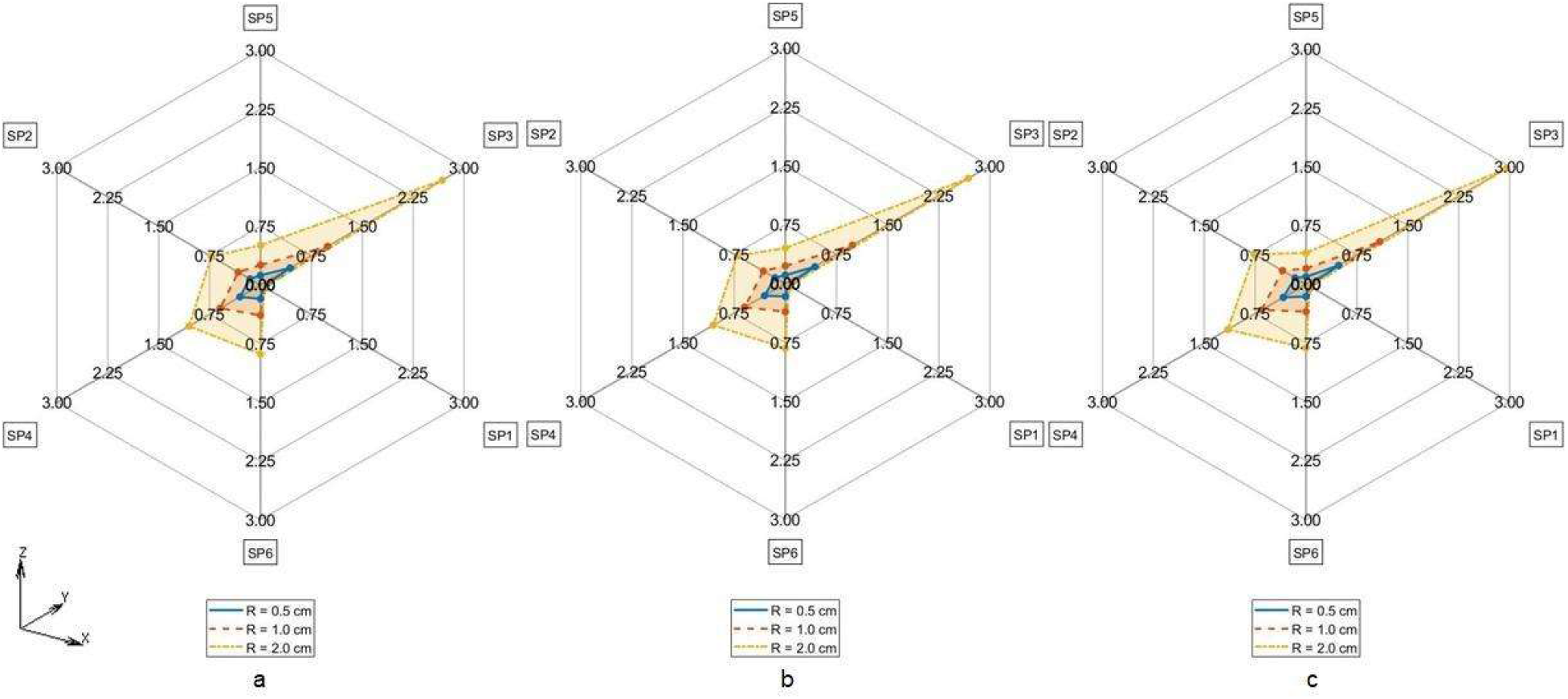
TDE deviation in D_MAX_ for CTM for R = 0.5 cm (Solid line), R = 1.0 cm (Dashed line), R = 2 cm (Dash-dotted line) a. T_A_ = 5° C, b. T_A_ = 15° C, c. T_A_ = 25° C

**Fig. 6:**
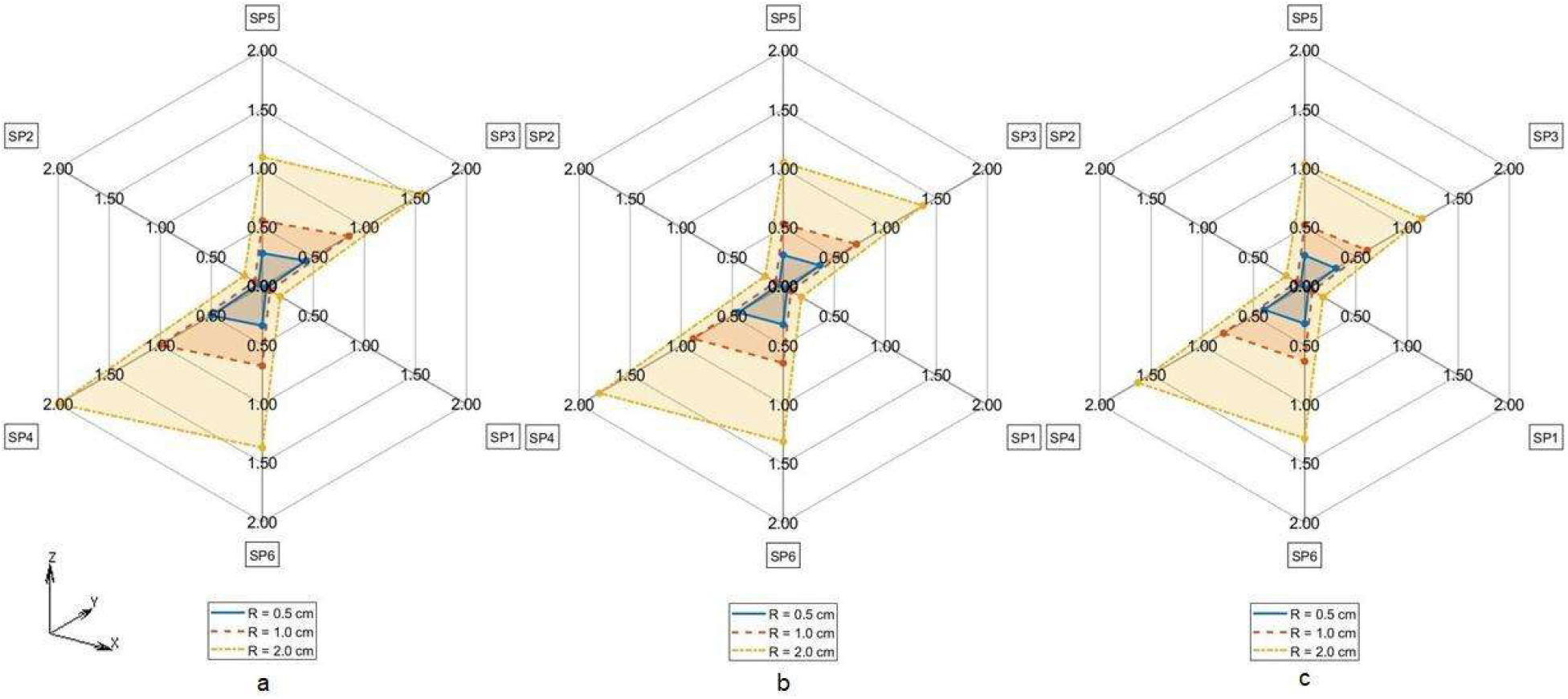
TDE deviation in D_MAX_ for CMS for R = 0.5 cm (Solid line), R = 1.0 cm (Dashed line), R = 2 cm (Dash-dotted line) a. T_A_ = 5° C, b. T_A_ = 15° C, c. T_A_ = 25° C

**Fig. 7:**
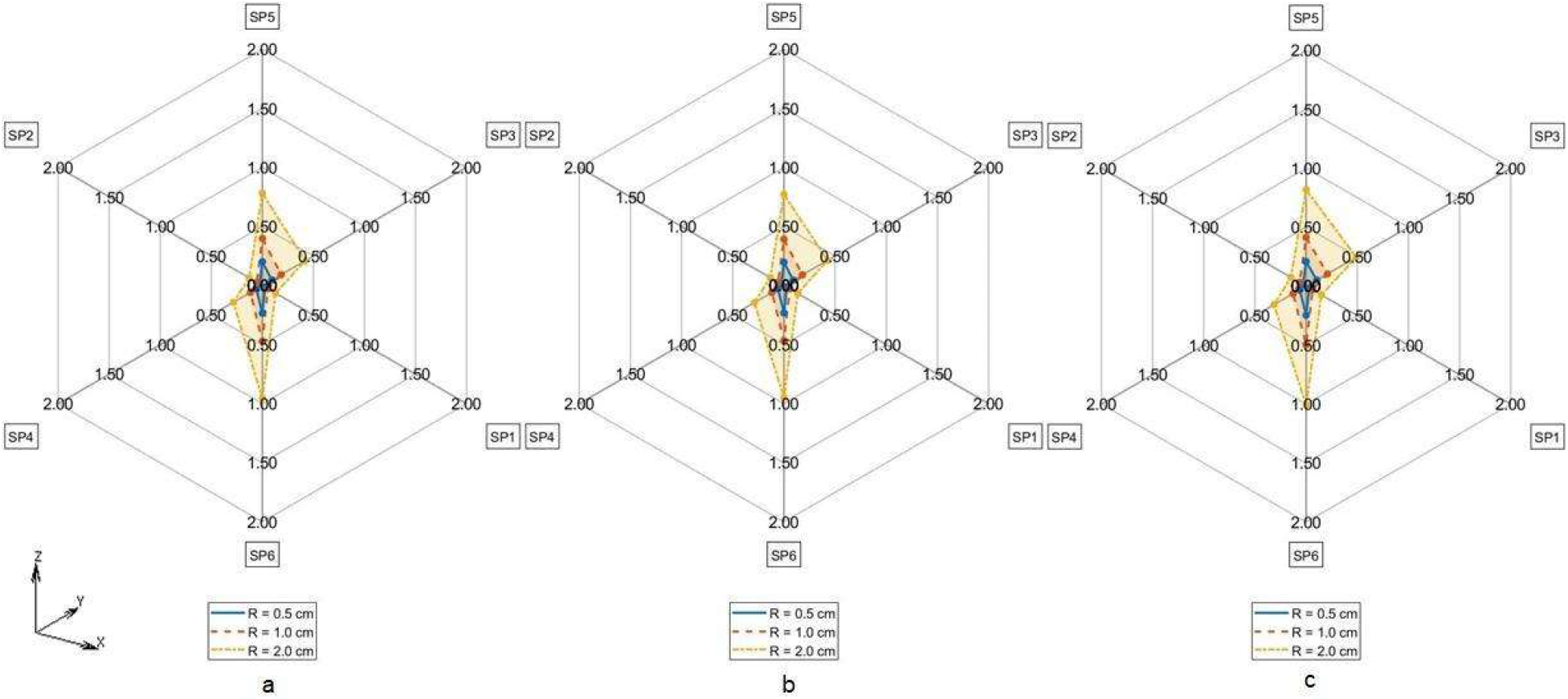
TDE deviation in D_MAX_ for CM for R = 0.5 cm (Solid line), R = 1.0 cm (Dashed line), R = 2 cm (Dash-dotted line) a. T_A_ = 5° C, b. T_A_ = 15° C, c. T_A_ = 25° C

**Fig. 8:**
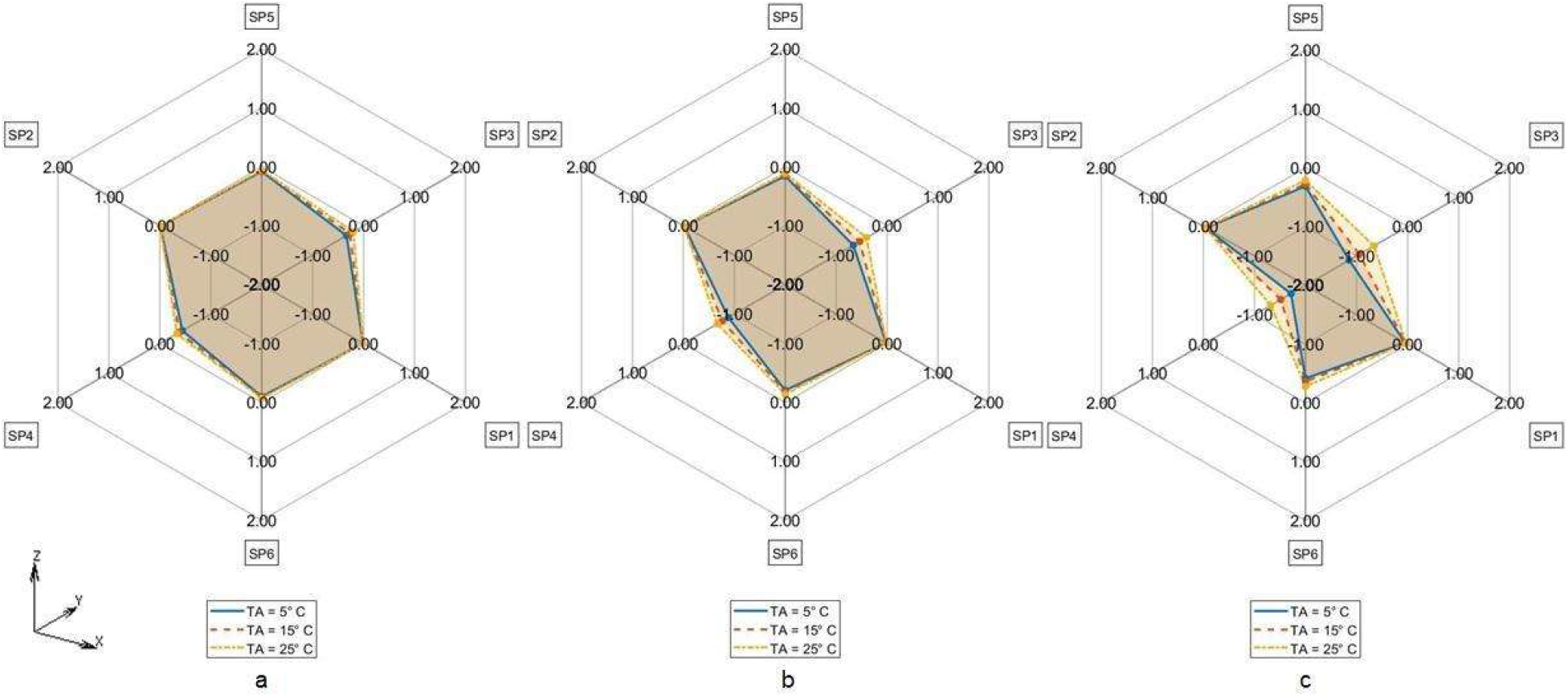
Difference in TDE deviation in D_MAX_ between CM vs CMS model for T_A_ = 5° C (Solid line), T_A_ = 15° C (Dashed line), T_A_ = 25° C (Dash-dotted line) for a. R = 0.5 cm, b. R = 1 cm, c. R = 2 cm

TDE deviations D_MAX,Qi_^M^ at various measurement points SP_R_k (k = 1, …, 6) evaluated against C in M = CTM, CMS, CM at various ambient temperatures T_A_ and Q regions are shown in Tab. 2 – Tab. 10 in the Appendix. The RTML SP_R_k (k = 1, …, 6) in Tab. 2 – Tab. 10 are arranged corresponding to clockwise order in Fig. 5 – Fig. 9. Browsing the tables the following regularities can be noticed:

**Tab. 2:** TDE deviation D_MAX_^M^ in M = CTM at T_A_ = 5° C. Maximum value for fixed R and fixed SP_R_k: light grey, maximum value for fixed R over all SP_R_k: fat print, dark grey

| TDE deviation – CTM model - $D_{MAX}$ | | | | | | |
| --- | --- | --- | --- | --- | --- | --- |
| TA = 5° C, R = 0.5 cm |  |  |  |  |  |  |
|  | SP5 | SP3 | SP1 | SP6 | SP4 | SP2 |
| Q1 | 0.122852 | 0.093391 | 0.072357 | 0.055339 | **0.435910** | 0.386072 |
| Q2 | 0.373923 | 0.369764 | 0.044560 | 0.038094 | 0.034864 | 0.032667 |
| Q3 | 0.176460 | 0.141712 | 0.120125 | 0.103497 | 0.303744 | 0.270017 |
| Q4 | 0.262275 | 0.260607 | 0.155169 | 0.134974 | 0.127751 | 0.123630 |
| TA = 5° C, R = 1 cm |  |  |  |  |  |  |
| Q1 | 0.255475 | 0.191303 | 0.145863 | 0.109616 | **0.989255** | 0.875922 |
| Q2 | 0.843773 | 0.833404 | 0.070145 | 0.059609 | 0.054310 | 0.050870 |
| Q3 | 0.387430 | 0.311325 | 0.265372 | 0.231326 | 0.593254 | 0.527706 |
| Q4 | 0.511466 | 0.507438 | 0.329621 | 0.286434 | 0.270129 | 0.261497 |
| TA = 5° C, R = 2 cm |  |  |  |  |  |  |
| Q1 | 0.508475 | 0.368379 | 0.273719 | 0.200823 | **2.676332** | 2.381748 |
| Q2 | 2.272005 | 2.236726 | 0.055936 | 0.045853 | 0.040050 | 0.036235 |
| Q3 | 0.884038 | 0.707707 | 0.604823 | 0.533477 | 1.038491 | 0.925452 |
| Q4 | 0.898327 | 0.891724 | 0.724933 | 0.628021 | 0.586598 | 0.566291 |

**Tab. 3:**
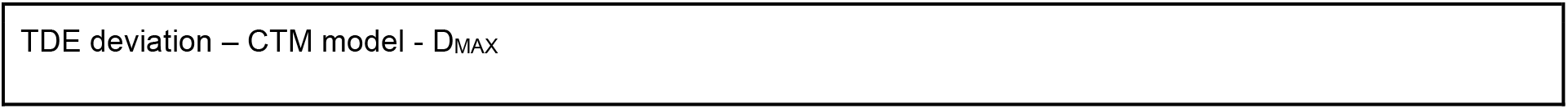
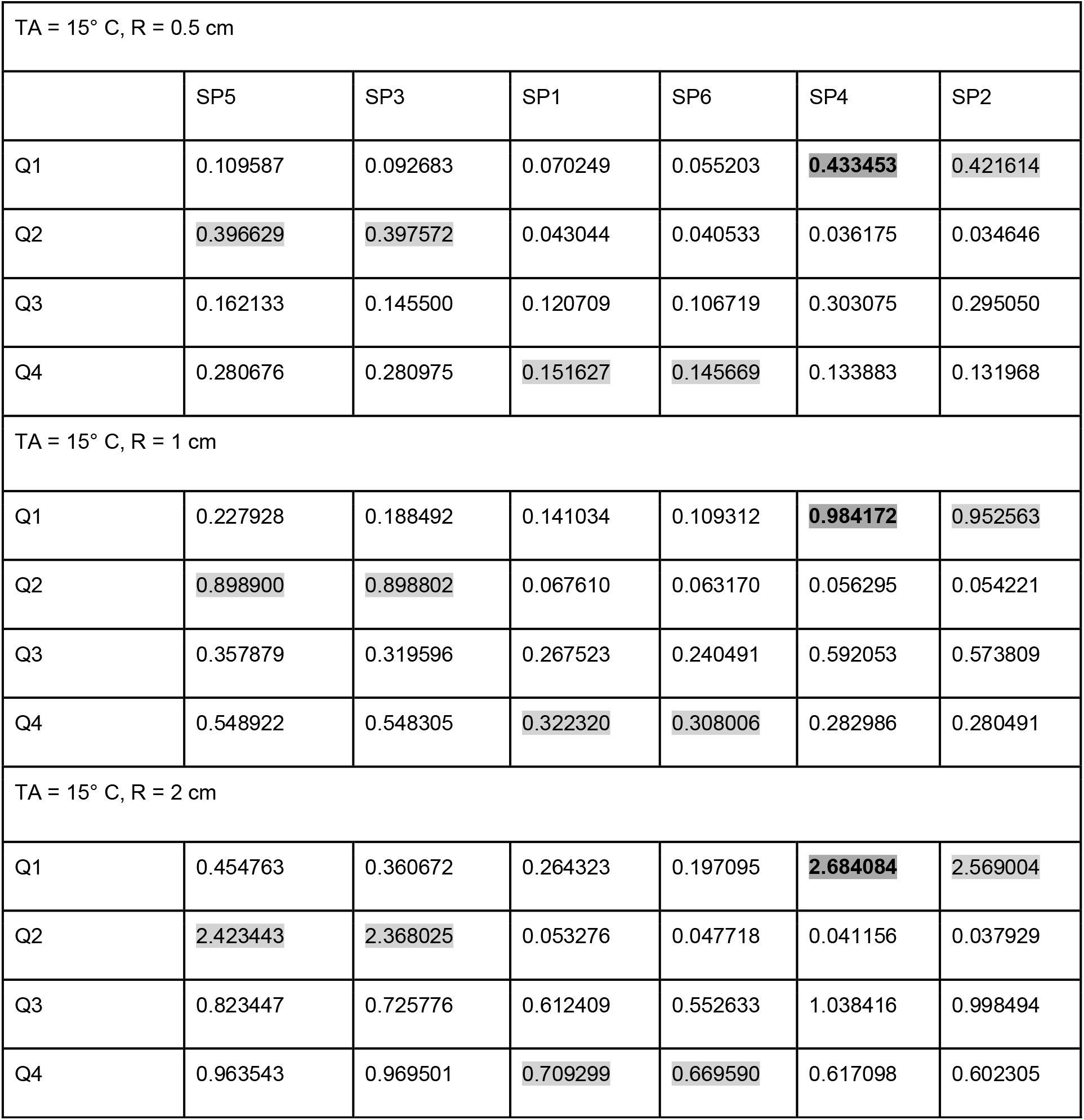
TDE deviation D_MAX_^M^ in M = CTM at T_A_ = 15° C. Maximum value for fixed R and fixed SP_R_k: light grey, maximum value for fixed R over all SP_R_k: fat print, dark grey

| TDE deviation – CTM model - $D_{MAX}$ |
| --- |

**Tab. 3:** TDE deviation $D_{MAX}^M$ in $M = CTM$ at $T_A = 15^\circ C$ . Maximum value for fixed R and fixed $SP_{Rk}$ : light grey, maximum value for fixed R over all $SP_{Rk}$ : fat print, dark grey
|  |  |  |  |  |  |  |
| --- | --- | --- | --- | --- | --- | --- |
| TA = 15° C, R = 0.5 cm |  |  |  |  |  |  |
|  | SP5 | SP3 | SP1 | SP6 | SP4 | SP2 |
| Q1 | 0.109587 | 0.092683 | 0.070249 | 0.055203 | **0.433453** | 0.421614 |
| Q2 | 0.396629 | 0.397572 | 0.043044 | 0.040533 | 0.036175 | 0.034646 |
| Q3 | 0.162133 | 0.145500 | 0.120709 | 0.106719 | 0.303075 | 0.295050 |
| Q4 | 0.280676 | 0.280975 | 0.151627 | 0.145669 | 0.133883 | 0.131968 |
| TA = 15° C, R = 1 cm |  |  |  |  |  |  |
| Q1 | 0.227928 | 0.188492 | 0.141034 | 0.109312 | **0.984172** | 0.952563 |
| Q2 | 0.898900 | 0.898802 | 0.067610 | 0.063170 | 0.056295 | 0.054221 |
| Q3 | 0.357879 | 0.319596 | 0.267523 | 0.240491 | 0.592053 | 0.573809 |
| Q4 | 0.548922 | 0.548305 | 0.322320 | 0.308006 | 0.282986 | 0.280491 |
| TA = 15° C, R = 2 cm |  |  |  |  |  |  |
| Q1 | 0.454763 | 0.360672 | 0.264323 | 0.197095 | **2.684084** | 2.569004 |
| Q2 | 2.423443 | 2.368025 | 0.053276 | 0.047718 | 0.041156 | 0.037929 |
| Q3 | 0.823447 | 0.725776 | 0.612409 | 0.552633 | 1.038416 | 0.998494 |
| Q4 | 0.963543 | 0.969501 | 0.709299 | 0.669590 | 0.617098 | 0.602305 |

**Tab. 4:** TDE deviation D_MAX_^M^ in M = CTM at T_A_ = 25° C. Maximum value for fixed R and fixed SP_R_k: light grey, maximum value for fixed R over all SP_R_k: fat print, dark grey

|  |  |  |  |  |  |  |
| --- | --- | --- | --- | --- | --- | --- |
| TDE deviation – CTM model - $D_{MAX}$ | | | | | | |
| TA = 25° C, R = 0.5 cm |  |  |  |  |  |  |
|  | SP5 | SP3 | SP1 | SP6 | SP4 | SP2 |
| Q1 | 0.096924 | 0.087224 | 0.064009 | 0.047629 | **0.478770** | 0.472093 |
| Q2 | 0.433839 | 0.420362 | 0.044949 | 0.043581 | 0.038244 | 0.035446 |
| Q3 | 0.155922 | 0.146795 | 0.119631 | 0.102337 | 0.336467 | 0.331520 |
| Q4 | 0.305825 | 0.297609 | 0.163489 | 0.160196 | 0.144541 | 0.137588 |
| TA = 25° C, R = 1 cm |  |  |  |  |  |  |
| Q1 | 0.200676 | 0.176616 | 0.127789 | 0.093306 | **1.085921** | 1.064973 |
| Q2 | 0.978343 | 0.949218 | 0.070626 | 0.067887 | 0.059601 | 0.055385 |
| Q3 | 0.348042 | 0.325264 | 0.267879 | 0.232388 | 0.657130 | 0.643892 |
| Q4 | 0.594666 | 0.580189 | 0.347716 | 0.338657 | 0.305722 | 0.291640 |
| TA = 25° C, R = 2 cm |  |  |  |  |  |  |
| Q1 | 0.401715 | 0.336522 | 0.240250 | 0.171164 | **2.953641** | 2.854130 |
| Q2 | 2.631010 | 2.552613 | 0.054170 | 0.050111 | 0.042954 | 0.038911 |
| Q3 | 0.813108 | 0.743778 | 0.622384 | 0.548695 | 1.153148 | 1.114621 |
| Q4 | 1.035954 | 1.013757 | 0.765055 | 0.732501 | 0.663553 | 0.634064 |

**Tab. 5:** TDE deviation D_MAX_^M^ in M = CMS at T_A_ = 5° C. Maximum value for fixed R and fixed SP_R_k: light grey, maximum value for fixed R over all SP_R_k: fat print, dark grey

|  |  |  |  |  |  |  |
| --- | --- | --- | --- | --- | --- | --- |
| TDE deviation – CMS model - $D_{MAX}$ | | | | | | |
| TA = 5° C, R = 0.5 cm |  |  |  |  |  |  |
|  | SP5 | SP3 | SP1 | SP6 | SP4 | SP2 |
| Q1 | 0.278668 | 0.428958 | 0.036924 | 0.335980 | **0.499407** | 0.036924 |
| Q2 | 0.237840 | 0.249328 | 0.034020 | 0.287842 | 0.348655 | 0.034020 |
| Q3 | 0.187893 | 0.116238 | 0.029856 | 0.228624 | 0.222521 | 0.029856 |
| Q4 | 0.154688 | 0.074618 | 0.027514 | 0.190183 | 0.145349 | 0.027514 |
| TA = 5° C, R = 1 cm |  |  |  |  |  |  |
| Q1 | 0.554121 | 0.847941 | 0.073925 | 0.676501 | **1.002985** | 0.073925 |
| Q2 | 0.459769 | 0.466711 | 0.066970 | 0.564490 | 0.663992 | 0.066970 |
| Q3 | 0.363333 | 0.208421 | 0.059023 | 0.449124 | 0.421534 | 0.059023 |
| Q4 | 0.298674 | 0.161567 | 0.054472 | 0.373160 | 0.273236 | 0.054472 |
| TA = 5° C, R = 2 cm |  |  |  |  |  |  |
| Q1 | 1.095381 | 1.563284 | 0.176399 | 1.371045 | **1.999290** | 0.176399 |
| Q2 | 0.860177 | 0.756267 | 0.154756 | 1.086690 | 1.247146 | 0.154756 |
| Q3 | 0.682823 | 0.273074 | 0.137774 | 0.868890 | 0.800136 | 0.137774 |
| Q4 | 0.557965 | 0.424460 | 0.127253 | 0.719497 | 0.524725 | 0.127253 |

**Tab. 6:** TDE deviation D_MAX_^M^ in M = CMS at T_A_ = 15° C. Maximum value for fixed R and fixed SP_R_k: light grey, maximum value for fixed R over all SP_R_k: fat print, dark grey

|  |  |  |  |  |  |  |
| --- | --- | --- | --- | --- | --- | --- |
| TDE deviation – CMS model - $D_{MAX}$ | | | | | | |
| TA = 15° C, R = 0.5 cm |  |  |  |  |  |  |
|  | SP5 | SP3 | SP1 | SP6 | SP4 | SP2 |
| Q1 | 0.267230 | 0.357906 | 0.037319 | 0.322041 | **0.439618** | 0.037319 |
| Q2 | 0.247672 | 0.240927 | 0.036283 | 0.299441 | 0.353170 | 0.036283 |
| Q3 | 0.196644 | 0.114936 | 0.031731 | 0.239472 | 0.230824 | 0.031731 |
| Q4 | 0.158513 | 0.073921 | 0.028449 | 0.194800 | 0.149584 | 0.028449 |
| TA = 15° C, R = 1 cm |  |  |  |  |  |  |
| Q1 | 0.531431 | 0.717470 | 0.074719 | 0.648396 | **0.881242** | 0.074719 |
| Q2 | 0.482692 | 0.458454 | 0.071888 | 0.592171 | 0.682468 | 0.071888 |
| Q3 | 0.380752 | 0.209685 | 0.062621 | 0.471052 | 0.440656 | 0.062621 |
| Q4 | 0.307615 | 0.158411 | 0.056403 | 0.384086 | 0.284623 | 0.056403 |
| TA = 15° C, R = 2 cm |  |  |  |  |  |  |
| Q1 | 1.050738 | 1.369435 | 0.178258 | 1.313951 | **1.804971** | 0.178258 |
| Q2 | 0.915332 | 0.762669 | 0.167574 | 1.155611 | 1.314195 | 0.167574 |
| Q3 | 0.715042 | 0.285432 | 0.145382 | 0.911878 | 0.848165 | 0.145382 |
| Q4 | 0.579908 | 0.414947 | 0.132116 | 0.746680 | 0.559106 | 0.132116 |

**Tab. 7:** TDE deviation D_MAX_^M^ in M = CMS at T_A_ = 25° C. Maximum value for fixed R and fixed SP_R_k: light grey, maximum value for fixed R over all SP_R_k: fat print, dark grey

|  |  |  |  |  |  |  |
| --- | --- | --- | --- | --- | --- | --- |
| TDE deviation – CMS model - $D_{MAX}$ | | | | | | |
| TA = 25° C, R = 0.5 cm |  |  |  |  |  |  |
|  | SP5 | SP3 | SP1 | SP6 | SP4 | SP2 |
| Q1 | 0.262335 | 0.305714 | 0.037699 | 0.315216 | **0.395636** | 0.037699 |
| Q2 | 0.246257 | 0.194035 | 0.036956 | 0.296862 | 0.319504 | 0.036956 |
| Q3 | 0.201215 | 0.085054 | 0.033200 | 0.244457 | 0.214002 | 0.033200 |
| Q4 | 0.168303 | 0.085014 | 0.030656 | 0.206545 | 0.144324 | 0.030656 |
| TA = 25° C, R = 1 cm |  |  |  |  |  |  |
| Q1 | 0.521653 | 0.610921 | 0.075482 | 0.634763 | 0.792969 | 0.075482 |
| Q2 | 0.482290 | 0.371160 | 0.073451 | 0.589914 | 0.621087 | 0.073451 |
| Q3 | 0.393083 | 0.155605 | 0.065927 | 0.484524 | 0.413476 | 0.065927 |
| Q4 | 0.327041 | 0.179284 | 0.060712 | 0.408028 | 0.276591 | 0.060712 |
| TA = 25° C, R = 2 cm |  |  |  |  |  |  |
| Q1 | 1.031287 | 1.148291 | 0.180043 | 1.286870 | 1.631598 | 0.180043 |
| Q2 | 0.923440 | 0.610350 | 0.172432 | 1.163235 | 1.221785 | 0.172432 |
| Q3 | 0.751407 | 0.193555 | 0.154966 | 0.953344 | 0.821985 | 0.154966 |
| Q4 | 0.617182 | 0.448666 | 0.141694 | 0.796183 | 0.555555 | 0.141694 |

**Tab. 8:**
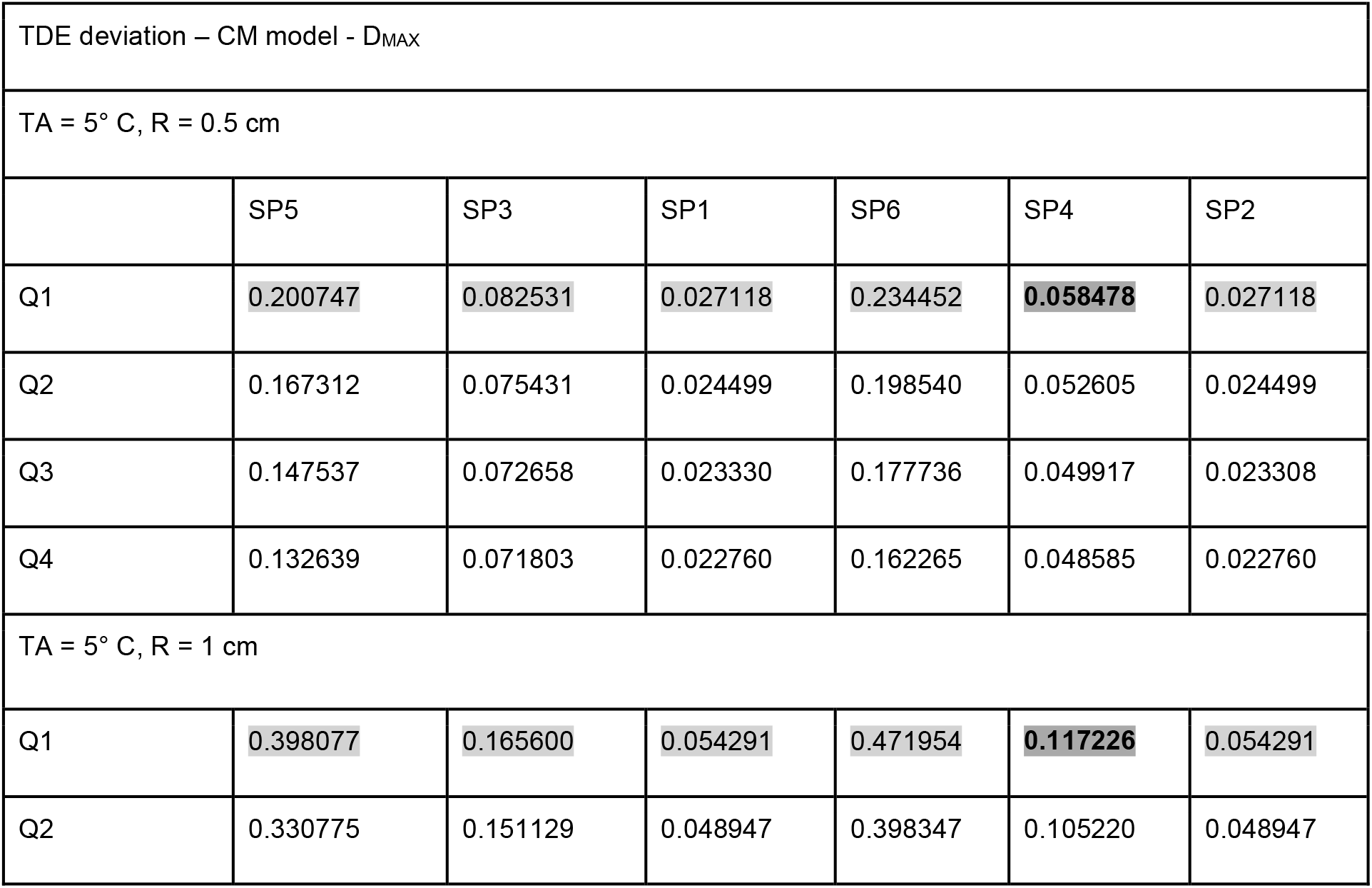
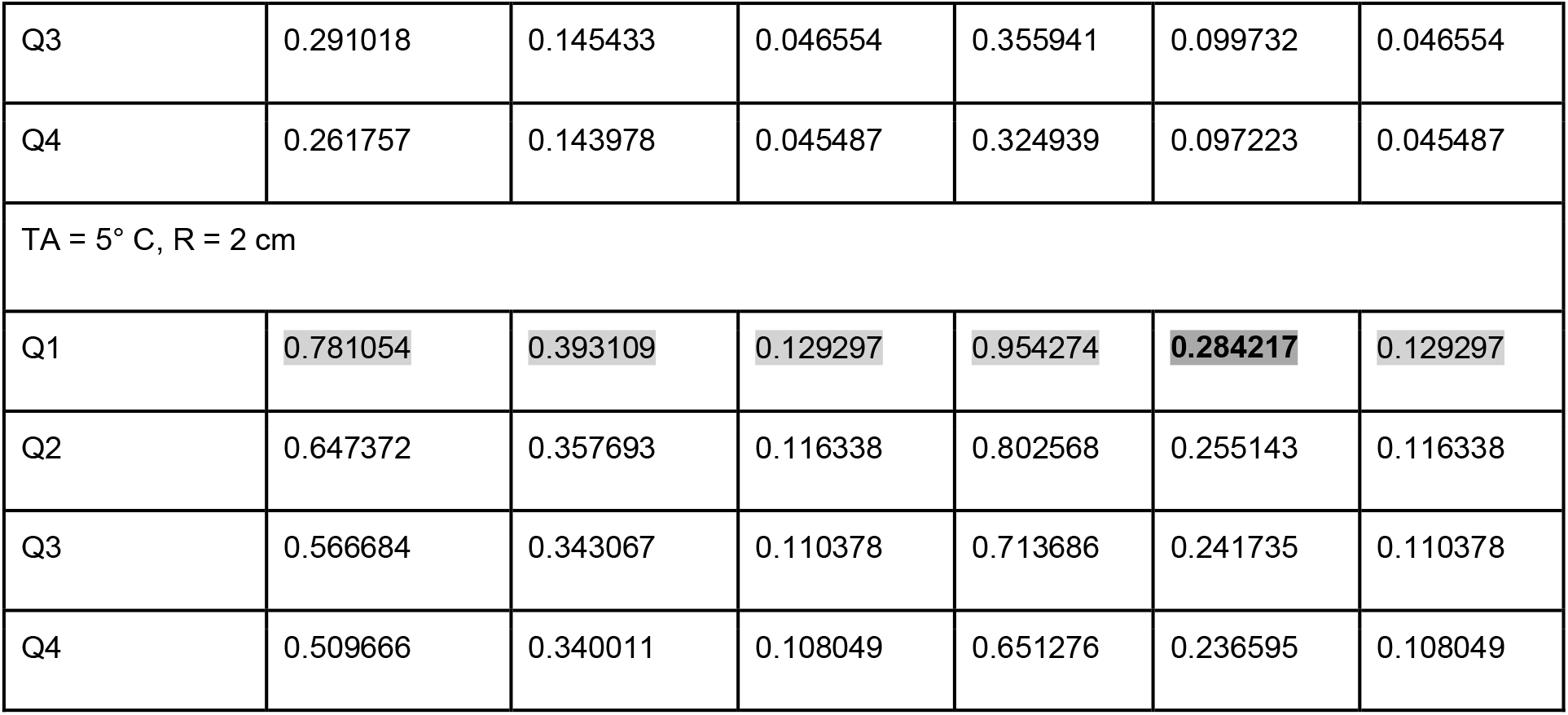
TDE deviation D_MAX_^M^ in M = CM at T_A_ = 5° C. Maximum value for fixed R and fixed SP_R_k: light grey, maximum value for fixed R over all SP_R_k: fat print, dark grey

**Tab. 9:**
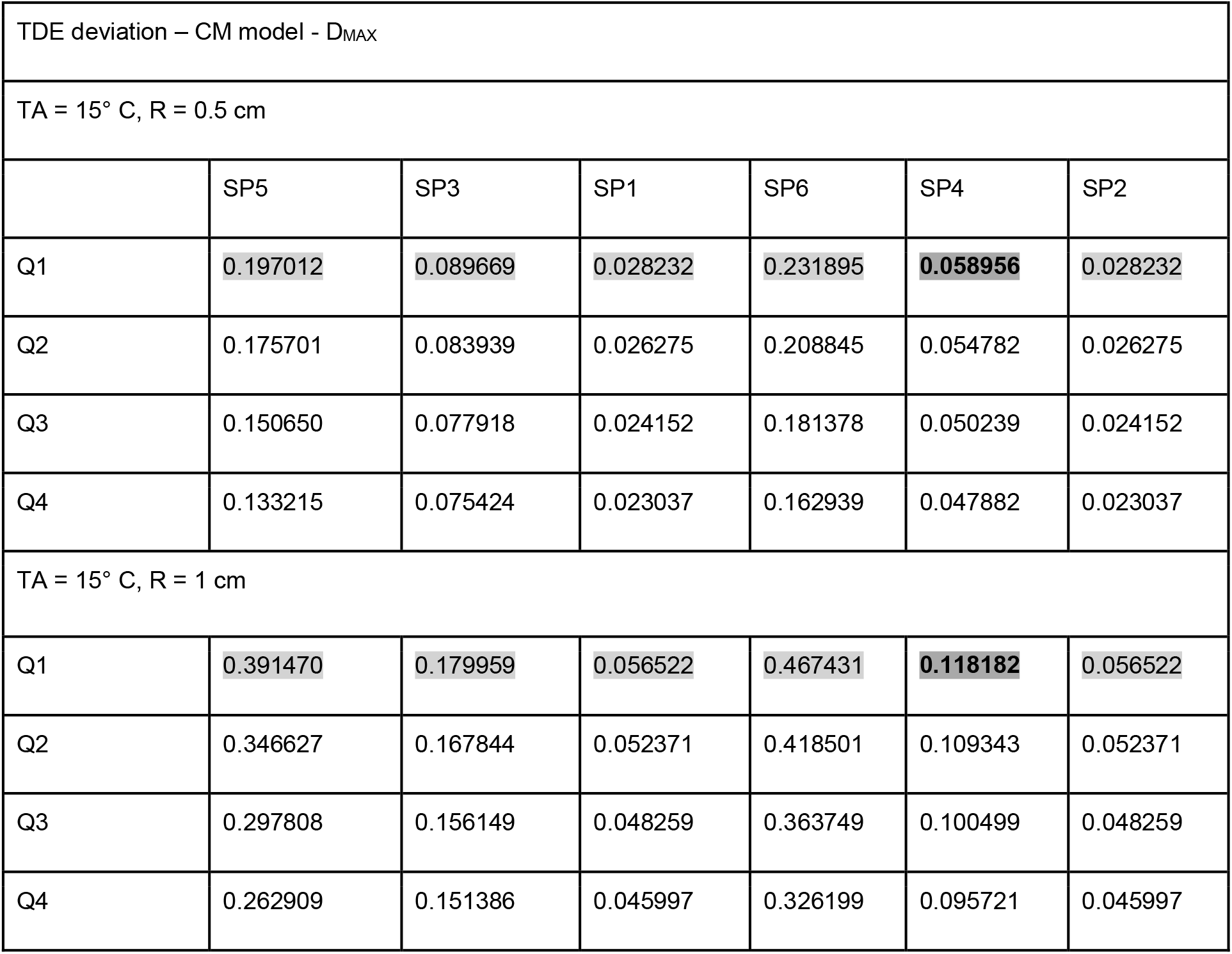
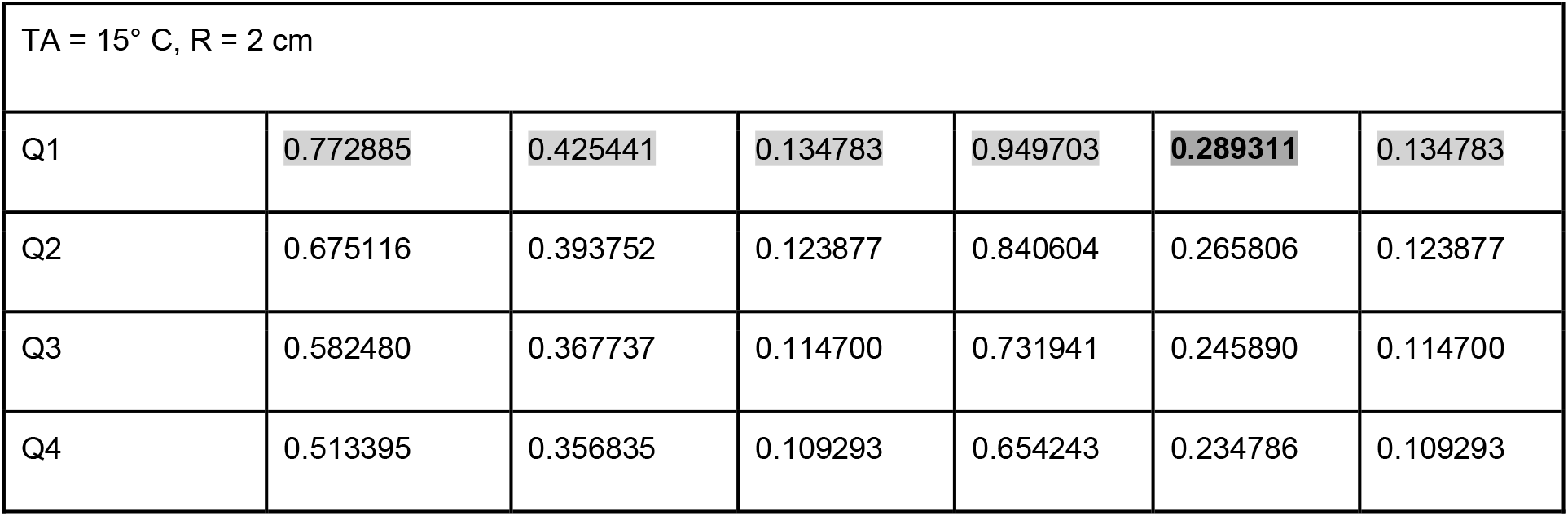
TDE deviation D_MAX_^M^ in M = CM at T_A_ = 15° C. Maximum value for fixed R and fixed SP_R_k: light grey, maximum value for fixed R over all SP_R_k: fat print, dark grey

**Tab. 10:**
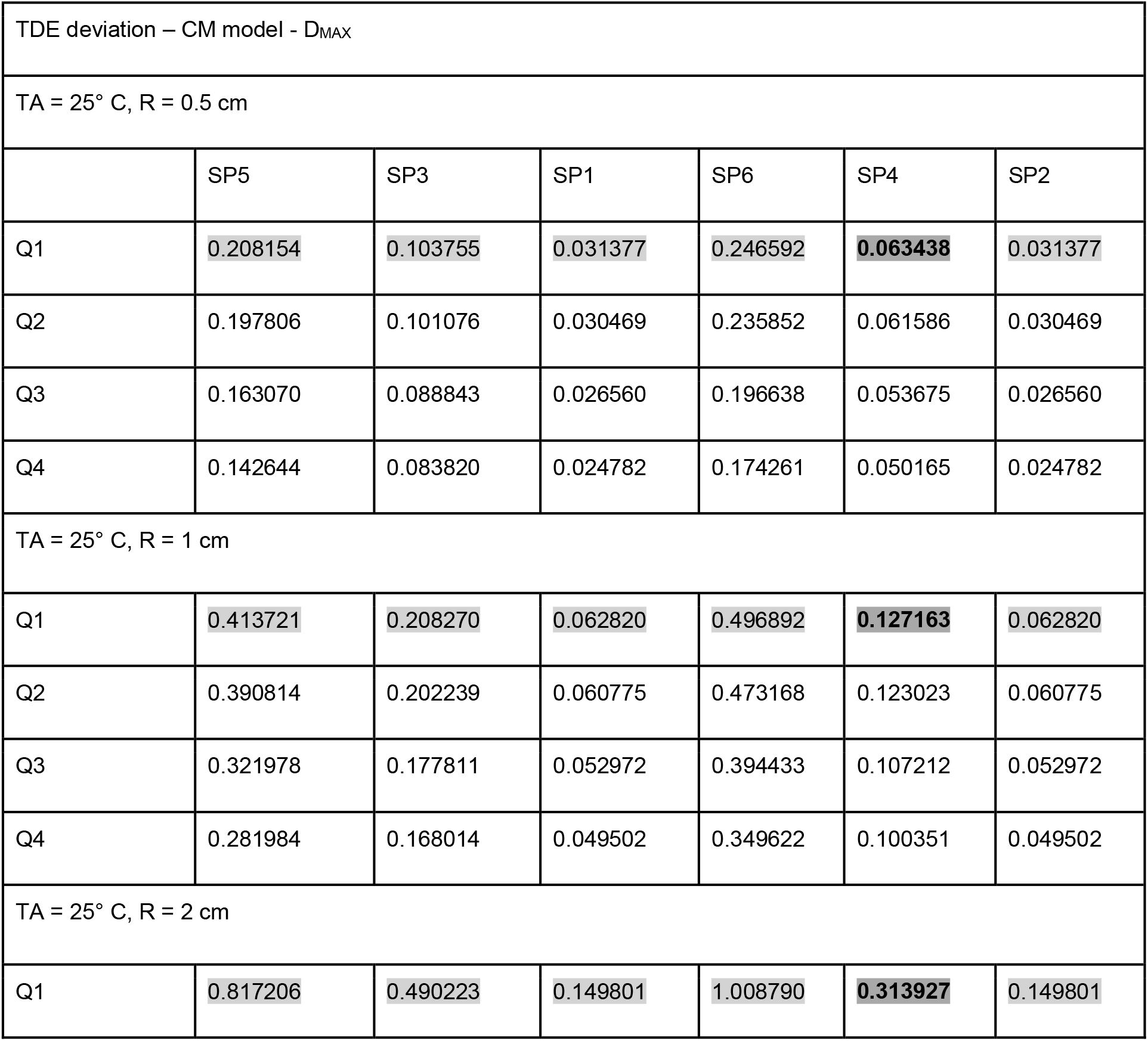
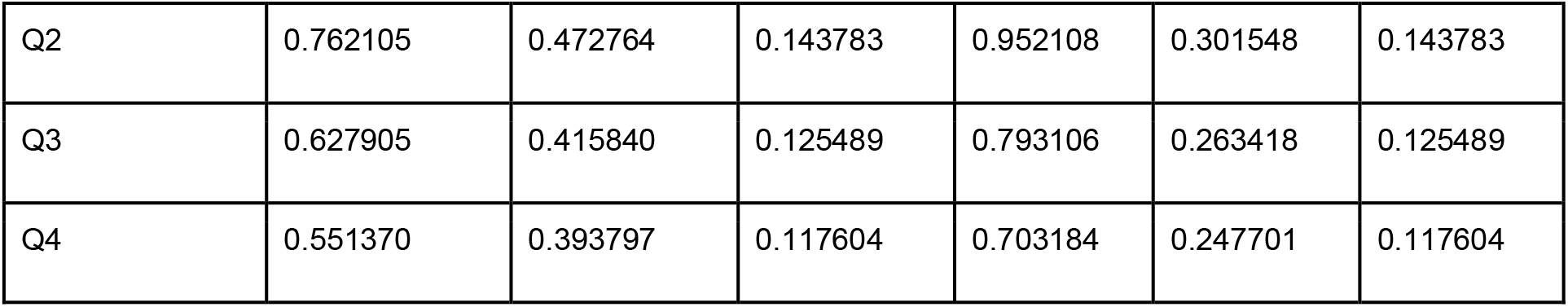
TDE deviation D_MAX_^M^ in M = CM at T_A_ = 25° C. Maximum value for fixed R and fixed SP_R_k: light grey, maximum value for fixed R over all SP_R_k: fat print, dark grey

**Fig. 9:**
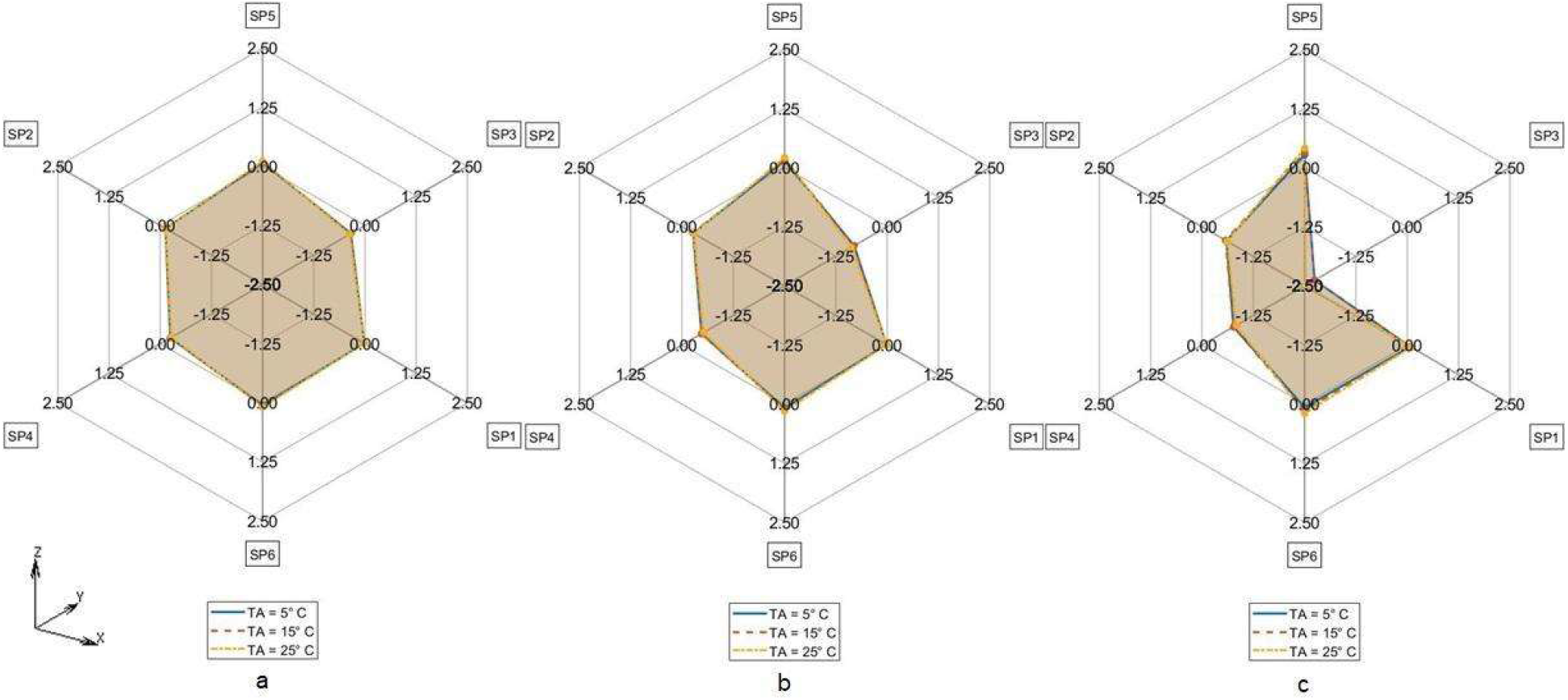
Difference in TDE deviation in D_MAX_ between CM vs CTM for T_A_ = 5° C (Solid line), T_A_ = 15° C (Dashed line), T_A_ = 25° C (Dash-dotted line) a. R = 0.5 cm, b. R = 1 cm, c. R = 2 cm

(R1) For all of the models M = CTM, CMS, CM, and for each fixed combination of T_A_ and R the maximum value of D_MAX,Qi_ over all Qi and over all SP_R_k is taken at SP_R_4 in Q1 (dark grey, fat print in Tab. 2 - 10).

(R2) For M = CM, CMS, and for each fixed combination of T_A_, R, SP_R_k the maximum value of D_MAX,Qi_ over all Qi is taken in Q1 (light grey in Tab. 5 – 10).

(R3) For M = CTM, and for each fixed combination of T_A_, R, the maxima of D_MAX,Qi_ for fixed SP_R_k are as follows:

a. In the SP_R_5- and in the SP_R_3-column at Q2
b. In the SP_R_1- and in the SP_R_6-column at Q4
c. In the SP_R_2-column at Q1

(R4) For M = CM, CMS, and for each fixed combination of T_A_, R, SP_R_k the D_MAX,Qi_ values are declining with rising index i of Q_i_. The only exception occurs for M = CMS in Tab. 7 and T_A_ = 25 °C at R = 1 cm and R = 2 cm at SP_R_3: where we see D_MAX,Qi_ rise from Q3 to Q4

Results concerning relative TDE difference distances and distances based on the L_2_-norm instead of the MAX norm are presented in the electronic SI.

## Discussion

Multiple analysis was carried out to determine the TDE error caused by measurement locus variations for the different FE-model types such as CTM, CM, CMS using maximum distance D_MAX_.

Our simulation results show maximum TDE deviations D_MAX_ with respect to variations in RTML up to 2 cm in the order of magnitude of 2 to 3 hours. Comparing to other influences like variations in ambient temperature T_A_ (see [6], [7]), initial body core temperature T_0_ (see [20]), etc. the TDE deviations of 2 to 3 hours are not negligible and can cause considerable errors in TDE. Our RTML-caused TDE deviation results D_MAX_ lie within the 95% confidence interval of Henßge [15]. For the first Q interval, Henßge gives a value of 2.8 hours for standard as well as for non-standard conditions, which is in the same order of our TDE deviations caused by variations of the RTML.

The maximum TDE deviation D_MAX_ in CTM and in CMS is observed in the dorsal-ventral axis Y (Fig. 5: SP3, Fig. 6: SP4).

This may be due to the close vicinity of RTML to backbone tissue. A higher value of thermal conductivity of bone tissue of k = 0.75 W/m^2^K compared to the conductivity of other soft tissue types, will result in higher temperature gradient in the line C – nearest backbone parts. Similarly, a higher temperature gradient occurs along the Y: dorsal-ventral SP3 - C - SP4 axis in case of CMS due to high conductivity of substrate. Only in CM the axis SP6 – C – SP5 (Z: caudal - cranial) shows the highest D_MAX_ values (see Fig. 7). This may be the case because the CM model lacks the high conductivity substrate the CMS model contains. Other differences between CM and the other models might as well be part of the explanation: The CM shows symmetric anatomical structures with respect to the mid sagittal plane while the CTM does not. Furthermore, the CTM has a much finer discretization than the CM and a varying spatial tissue distribution due to the manual versus the CT-based mesh generation in CM and CTM respectively. Concerning the FE-meshwidth, we found only a minor effect on the TDE difference results as well as [7]. Hence, the TDE differences between the CM and the CTM can hardly be explained from differences in the discretization order.

The only difference between CMS and CM is that in CMS, the coarse-meshed human model lies on a discretized structure with wet soil’s thermal properties whereas CM consists of a coarse-meshed human model floating freely (like CTM model) in air without contact to other solid structures. The difference D_MAX_^CM,CMS^ compares CM vs. CMS model thus considering the influence of the support structure on the sensitivity of TDE against RTML variation. Like in forensic scenarios heat transfer from the corpse to support with high conductivity and / or heat capacity will increase heat flux through the models support-contact faces. Assuming a support with comparably low thermal conductivity, like a highly insulated mattress, a decrease in heat flux will be the consequence. The influence of different support materials on TDE was investigated in [21, 24]. Thermal properties of the support were also considered in the Henßge model, where supports with high thermal conductivities are taken into account by increasing the correction factor and vice versa (see e.g. [15]). Our results show that the influence of different supports on TDE sensitivity with respect to RTML variations need to be considered because they may lead to differences in temperature gradients which transform to differences in TDE.

The ambient temperature T_A_ influences the TDE sensitivity against RTML variations in the CMS model in such a way that the TDE deviation increases with decrease in T_A_. Possibly, this effect is due to the presence of the support structure along the dorsal-ventral axis (Y axis (SP4 – C – SP3)), which increases the rate of heat transfer. The initial temperature of the support structure was always set to the ambient temperature T_A_. In contrast, looking at the TDE difference D_MAX_^CM,CTM^ between D_MAX_ in CM and in CTM, there is nearly no influence of ambient temperature T_A_. Apparently, the influence of T_A_ on TDE difference is more profound when the support structure is present.

Concerning the influence of RTML variation radius R, the maximum TDE deviation D_MAX_ increases with increase in R in CTM, CMS and CM. This is due to greater Euclidean distance of measurement points from the actual measurement point, which is translated via temperature gradient to greater D_MAX_.

Fig. 10 gives the overview of the maximum TDE deviation by RTML variation evaluated by D_MAX_ for CTM, CMS and CM with respect to ambient temperature T_A_ and measurement variation radius R.

**Fig. 10:**
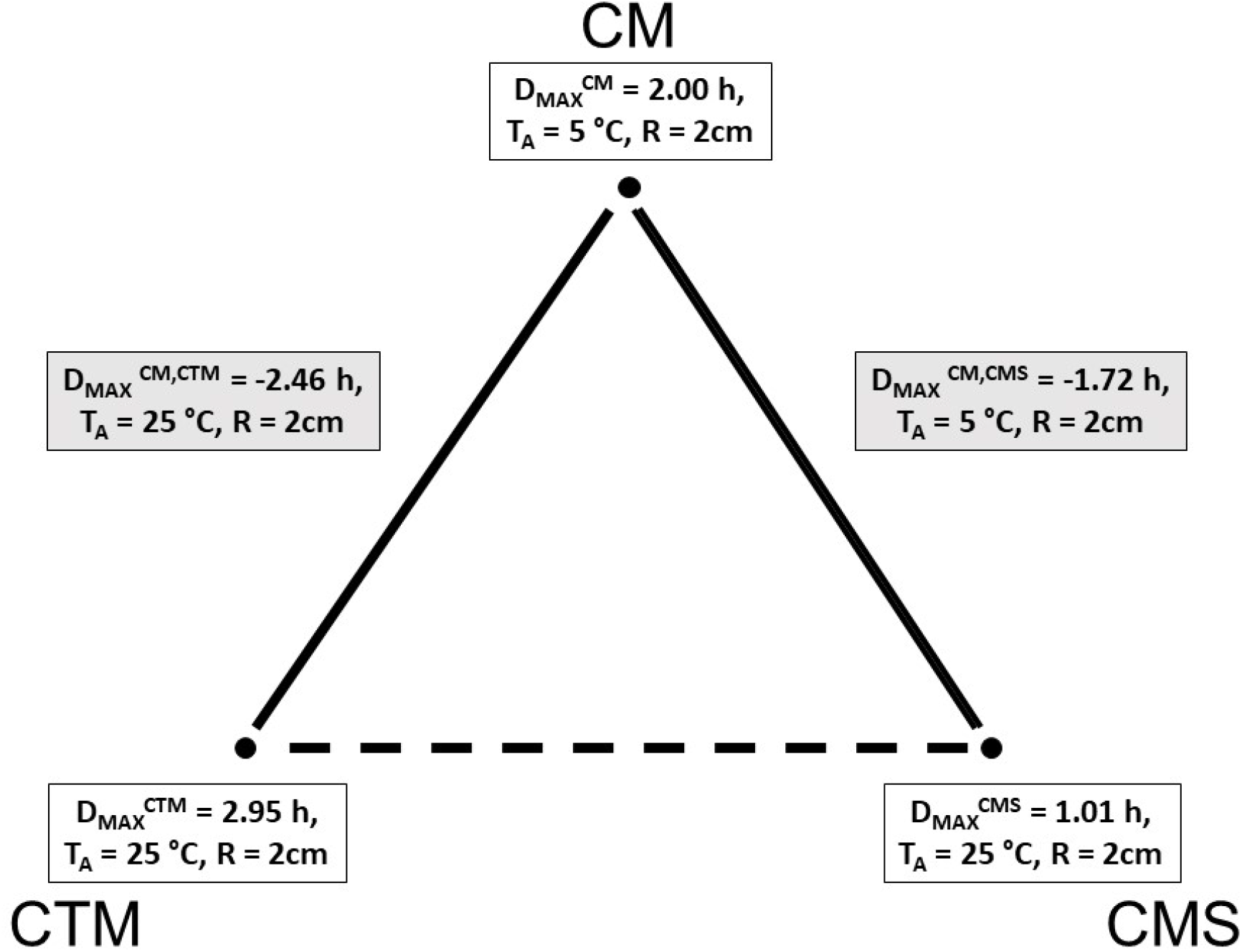
Maximum TDE deviation in D_MAX_ in CTM, CMS and CM model

The evaluation of the Qi-located TDE distances D_MAX,Qi_^M^ between SP_R_k and C yields the following results: Firstly measurement location errors in the ventral –dorsal axis Y seem to cause the most severe deviations of TDE (see (R1)). For the coarse model regardless of whether there is a wet soil substrate (M = CMS) or not (M = CM) the D_MAX,Qi_ values essentially decline with rising Q-index I that is for rising cooling times (see (R4)). Since the latter regularity is not repeated by the CT-generated model CTM, we infer this to be an artificial effect caused by the unrealistic high symmetry and regularity of the ‘hand-crafted’ FE models CMS and CM. Generally the effect of RTML errors on TDE seems to be highest at short times after death (see (R1)). Time evolution of TDE errors due to RTML variations thus correspond to the well known effect, that initial temperature deviations lead to high TDE errors during short times p.m. but are later damped to a low constant basic amount. This is shown in [6] for MHH – TDE but may easily be generalized. Moreover it is consistent with [7].

There are limitations of the current study. Firstly, we used a simulation-based approach for TDE estimation and RTML variation. A validation of our results on experimental measurements with real corpses was not realized. Measurements using corpses of recently deceased – aside from rare availability of such corpses - are difficult to carry out and especially a reproducible RTML variation is nearly impossible. However, in former studies the CM was calibrated and validated based on some experimental measurements published in [21]. Due to the physics-based approach, both the CM and the CTM should be suitable to carry out parameter variations in terms of RTML variation. Secondly, we only use three different models with normal body mass index. Cooling scenarios with more complex boundary conditions, like adipose corpses, clothing and coverings, etc. were not considered although these factors can influence TDE deviation as CMS depicts. Since varying all those boundary conditions would exceed the scope of one research article, those sensitivity studies are left for future research.

## Conclusions

TDE variations caused by RTML deviations may be a source of error in legal medicine TDE since they may reach a magnitude of 2-3 h. A possible consequence for legal medicine applied research could be to investigate methods and devices for reproducible RTML determination.

## Data Availability

All data produced in the present study are available upon reasonable request to the authors

## Acknowledgements

The present study is part of a project funded by the Deutsche Forschungsgemeinschaft DFG with the reference numbers MA 2501/4-1 (Prof. Dr. Gita Mall) and WE 2937/10-1 (Dr. Martin Weiser).

## Statements and Declarations

### Compliance with Ethical Standards

No living or deceased humans nor living or dead animals were directly involved in the study. Written consents of the CTM-subject’s kinship for the CT scan shown in the article were not needed since the body was confiscated by the local prosecution who directed the CT scan for investigations.

### Authors contributions

Study design and milestone discussions: All authors. CT-segmentation: ZIB authors. FE model CTM generation: ZIB authors, CTM migration: All authors. CMS generation: IRM authors. FE computations: J. Ulrich, J. Shanmugam, H. Muggenthaler. Distance measure computations: J. Shanmugam, M. Hubig. Output interpretation/presentation: J. Shanmugam, M. Hubig, H. Muggenthaler, J. Ulrich. Paper written/corrected: J. Shanmugam, M. Hubig, H. Muggenthaler. Paper reading / corrections: All authors.

### Material and/or code availability

The individual CT scan used in the current study is not publicly available. Each single slice is linked to a DICOM-file containing experiment parameter data and sensible case information. The program code is available from the corresponding author on reasonable request.

